# SARS-CoV-2 antibody responses in patients with aggressive haematological malignancies

**DOI:** 10.1101/2020.09.29.20202846

**Authors:** J. O’Nions, L. Muir, J. Zheng, C. Rees-Spear, A. Rosa, C. Earl, P. Cherepanov, R. Gupta, A. Khwaja, C. Jolly, L.E. McCoy

**Affiliations:** Department of Haematology, University College London NHS Foundation Trust, London, UK; Division of Infection and Immunity, University College London, London, UK; The Francis Crick Institute, London, UK; UCL Cancer Institute London, Paul O’Gorman Building, London, UK

## Abstract

The development of antibody responses to SARS-CoV-2 is an indicator of seroprevalence and may afford protection from infection. It has been presumed that antibody responses to SARS-CoV-2 will be impaired in patients with aggressive haematological malignancy (PHM) due to underlying immunological dysfunction caused by malignancy or systemic anti-cancer treatment (SACT), placing them at increased risk. Here we analysed longitudinal serum samples from ten hospitalised PHM with aggressive disease and on SACT, collected up to 103 days post-onset of COVID-19 symptoms. We found that the majority (8/9) of PHM with confirmed SARS-CoV-2 infection seroconverted and developed antibodies to the major SARS-CoV-2 antigens (S1 and N) with most (6/8) produced neutralising antibody responses. Furthermore, the dynamics of antibody responses were broadly similar to that reported for the general population, except for a possible delay to seroconversion. Our finding that PHM on SACT can make functional antibody responses to SARS-CoV-2 has important implications for patient management and serological monitoring of SARS-CoV-2 in high-risk groups.

## Introduction

COVID-19, the clinical disease caused by the novel coronavirus SARS-CoV-2, was first described in China in December 2019. Since then, it has developed into a global pandemic, with over 21.7 million people known to have been infected worldwide, and >41,000 deaths in the UK alone. Underlying cancer is a risk factor for poor prognosis and patients with haematological malignancy (PHM) are reported to have worst outcomes, although the prevalence, clinical course and outcomes of COVID-19 in PHM remain to be fully established^1, 2, 3, 4, 5, 6, 7^. Early reports of high mortality led to a number of recommendation to mitigate the impact of COVID-19 in PHM, including self-isolation and modification to standard therapies to reduce their immunosuppressive impact^8, 9^.

There is a significant global effort to understand antibody responses to SARS-CoV-2, not only to define cases and support immune-surveillance, but also to inform strategies for public health services and vaccination programs. Although the antibody response to SARS-CoV-2 in the general population is being increasingly characterised^10, 11^, reports in PHM are limited to a handful of cases reporting seropositivity^12, 13, 14^. The seroprevalence, magnitude, kinetics and functionality of the antibody response in in PHM remains unknown. Understanding this is critical as PHM are presumed to have impaired responses to SARS-CoV-2, due to either disease- or treatment-associated immune dysfunction. Here we investigate SARS-CoV-2 antibody responses in a cohort of patients with both aggressive haematological malignancies and COVID-19, who were receiving systemic anti-cancer therapy (SACT) at University College London Hospital during the pandemic.

## Methods

### Clinical cohort

Excess serum samples were stored for 10 patients with aggressive haematological malignancies and COVID-19 between April and May 2020. All clinical parameters, radiological and laboratory investigations were performed and recorded as part of routine standard of care.

### SARS-CoV-2 serological and neutralisation assays

Methods as described previously^15,16,17^

## Results and Discussion

We collected serial serum samples from 10 patients (A-K) who had COVID-19 and aggressive haematological malignancies (Table 1). Patients A-J tested positive for SARS-CoV-2 RNA by PCR. Patient K, was diagnosed with COVD-19 on clinical and radiological grounds, but was PCR-negative. Five patients had commenced SACT for haematological malignancy prior to contracting SARS-CoV-2 (median time from diagnosis of 30.5 days, IQR 106.5), five had *de novo* malignancies. Five patients had AML, three B-ALL, one T-ALL and one had primary CNS lymphoma (DLBCL-type). All patients received SACT within 28 days of developing COVID-19. Four patients received less myelosuppressive regimens (venetoclax azacitdine or gilteritinib) in accordance with NICE/NCRI COVID-19 guidance^9^. COVID-19 symptoms were assigned from mild to severe^15^, with two patients requiring ITU and mechanical ventilation. The median time from symptom onset and PCR diagnosis was 2 to all the patients who participated .5 days (IQR 26.5), the median duration of PCR positivity was 12 days (IQR 24) (Supplementary Fig. 1) and five patients received a potential COVID-19 modifying agent (tocilizumab, anakinra, remdesivir, hydroxychloroquine or dexamethasone). All patients survived and were discharged from hospital, with a median duration of illness of 22.5 days (IQR 32.25).

**Table 1:** Demographics, patient and disease characteristics, treatment and outcomes in patients with haematological malignancies and COVID-19.

| Patients (n=10) | A | B | C | D | E | F | G | H | J | K |
| --- | --- | --- | --- | --- | --- | --- | --- | --- | --- | --- |
| Age (years) | 45-49 | 25-29 | 35-39 | 20-24 | 60-64 | 50-54 | 65-69 | 35-39 | 55-59 | 75-79 |
| Sex | F | M | M | M | F | M | M | M | M | F |
| Ethnicity | Caucasian | Caucasian | Caucasian | South Asian | Caucasian | Caucasian | Middle Eastern | Caucasian | Caucasian | Caucasian |
| Haematological diagnosis | AML (new diagnosis) | AML (new diagnosis) | B ALL | AML | B ALL | T ALL | PCNSL/ DLBCL | AML | B ALL relapsed | AML |
| Haematological disease features | IDH2 mt | FLT3 mt; NPM1 WT | Normal CGN | Bi-allelic CEBPA mt, GATA2 mt | t(9;22), mono 7 | None | None | NPM1 mt, MECOM +1 | t(9;22) | Complex karyotype (del 5q, TP53 loss, mono 16, amplification KMT2A) |
| Haematological chemo- / immuno-therapy | Ven / Aza | AraC, Gilteritinib | Blina-tumumab (prev UKALL14) | DA | UKALL 60+ Ph+ induction | UKALL 14 Consolidation 1 | MATRIX | Ven / Aza | Mini FLA-Ida + imatinib, (prev UKALL14) | Ven / Aza |
| Days from haematological diagnosis to COVID-19 | 1 | 0 | 246 | 61 | 1 | 272 | 53 | 9 | 52 | 0 |
| Comorbidities | None | None | None | None | HTN T2DM | None | HTN T2DM CKD | None | None | COPD |
| Smoking history | Ex-smoker | None | None | None | None | None | None | Ex-smoker | Smoker | Ex-smoker |
| Presenting symptoms of COVID-19 | Fever, tooth abscess, myalgia, fatigue | Fever | Fever, collapse | Neutro-paenic fever | Cough, diarrhoea | Neutro-paenic fever | Fever, cough | Fever, cough | Fever | Fever, shortness of breath, palpitations |
| Days from symptom onset to COVID-19 | 35 | 3 | 2 | 0 | 28 | 1 | 0 | 3 | 1 | 27 |
| CXR / CT findings | None | Ground glass | Ground glass | Multiple areas of consolidation | Ground glass | None | None | Bilateral consolidation | Mild atelectasis | Ground glass |
| ITU admission | No | Yes | No | No | No | No | No | Yes | No | No |
| Max FiO2 | 21 | 100 | 85 | 21 | 21 | 21 | 36 | 100 | 24 | 60 |
| Max fever | 39.5 | 40.5 | 39.7 | 38.6 | 37.7 | 37.8 | 39.5 | 40.7 | 38 | 39.9 |
| COVID severity score* | 0 | 4 | 3 | 0 | 0 | 0 | 1 | 4 | 1 | 3 |
| **COVID-19 modifying treatment** | None | Dex | Anakinra | None | Remdesivir | None | HCQ | Tocilizumab | None | None |
| **Duration of PCR positivity (days)\*\*** | 8† | 59 | 33 | 11 | 8 | 12 | 9 | 32 | 25 | NA |
| **Days from symptom onset to negative PCR** | 43 | 62 | 35 | 11 | 36 | 13 | 9 | 35 | 26 | NA |
| **Duration of illness (days)\*\*\*** | 15 | 50 | 30 | 14 | 45 | 15 | 14 | 43 | 9 | 64 |
| **Outcome** | Alive, OP | Alive, OP | Alive, OP | Alive, OP | Alive, OP | Alive, OP | Alive, OP | Alive, OP | Alive, OP | Alive, OP |

\* COVID-19 severity score as previously defined<sup>15</sup>: 0 – asymptomatic OR no requirement for supplemental oxygen; 1 – supplemental oxygen ( $FiO_2 < 0.4$ ) for $\geq 12$ hours; 2 – supplemental oxygen ( $FiO_2 \geq 0.4$ ) for $\geq 12$ hours, 3 – requirement for NIV/CPAP OR proning OR supplemental oxygen ( $FiO_2 > 0.6$ ) for $\geq 12$ hours; 4 – intubation and ventilation OR supplemental oxygen ( $FiO_2 > 0.8$ ) AND peripheral $SpO_2 < 90\%$ (no known T2RF or $< 85\%$ if known T2RF) for $\geq 12$ .
\*\* RT PCR for SARS-CoV-2 was performed on samples from a combined nose and throat swab specimen.
\*\*\*Duration of illness defined as the period between diagnosis and cessation of treatment for COVID-19 that would mandate inpatient treatment (step down from ITU or discharge from the COVID ward).
† This patient subsequently tested positive one day after initial negative (for four days) again 21 days after second negative test (for eight days) (Supplementary Fig. 2)
All patients consented for excess serum to be stored and used as part of the “UCL Biobank for Studying Health and Disease – Haematology Project”, reference no NC10.13
Definitions: AML, acute myeloid leukaemia; B-ALL, B-lymphoblastic leukaemia; T-ALL, T lymphoblastic leukaemia; DLBCL, diffuse large B cell lymphoma; Ven / aza, venetoclax / azacytidine; DA, daunorubicin, AraC; MATRIX, methotrexate, rituximab, AraC; Dex, dexamethasone; HCQ, hydroxychloroquine; HTN, hypertension; CKD, chronic kidney disease; COPD, chronic obstructive pulmonary disease; T2DM, type 2 diabetes mellitus; OP, outpatient

Serum samples were taken between 2 and 103 days post onset of symptoms (POS), and a direct ELISA used to screen for anti-SARS-CoV-2 antibodies. Antibodies to the external Spike protein antigen were measured using a recombinant Spike S1_1-530_ subunit protein^10, 15^, and antibodies to the full-length internal Nucleoprotein (N) antigen measured to confirm infection^16^. A positive result was recorded when the sample absorbance was >4-fold above the average background of the assay (threshold established using >350 pre-2020 negative control sera as described previously^15, 16^). Total serum IgG was quantified and was in the normal range in each case, confirming that any reduction or absence of SARS-CoV-2 IgG was not due to hypogammaglubinaemia. Eight patients displayed detectable IgG responses to S1 and N (Fig. 1A,B and Supplementary Fig. 2). Two (F and K), generated neither S1 nor N antibody. K had a clinical diagnosis of COVID-19 but was PCR-negative. Interestingly, patient F (T-ALL) had clinically mild COVID-19 and did not receive COVID-19 modifying treatment. These results demonstrate that PHM can generate antibodies to SARS-CoV-2 antigens that are similar to those previously identified in COVID-19 patients without haematological malignancy^15, 16^.

**Figure 1:**
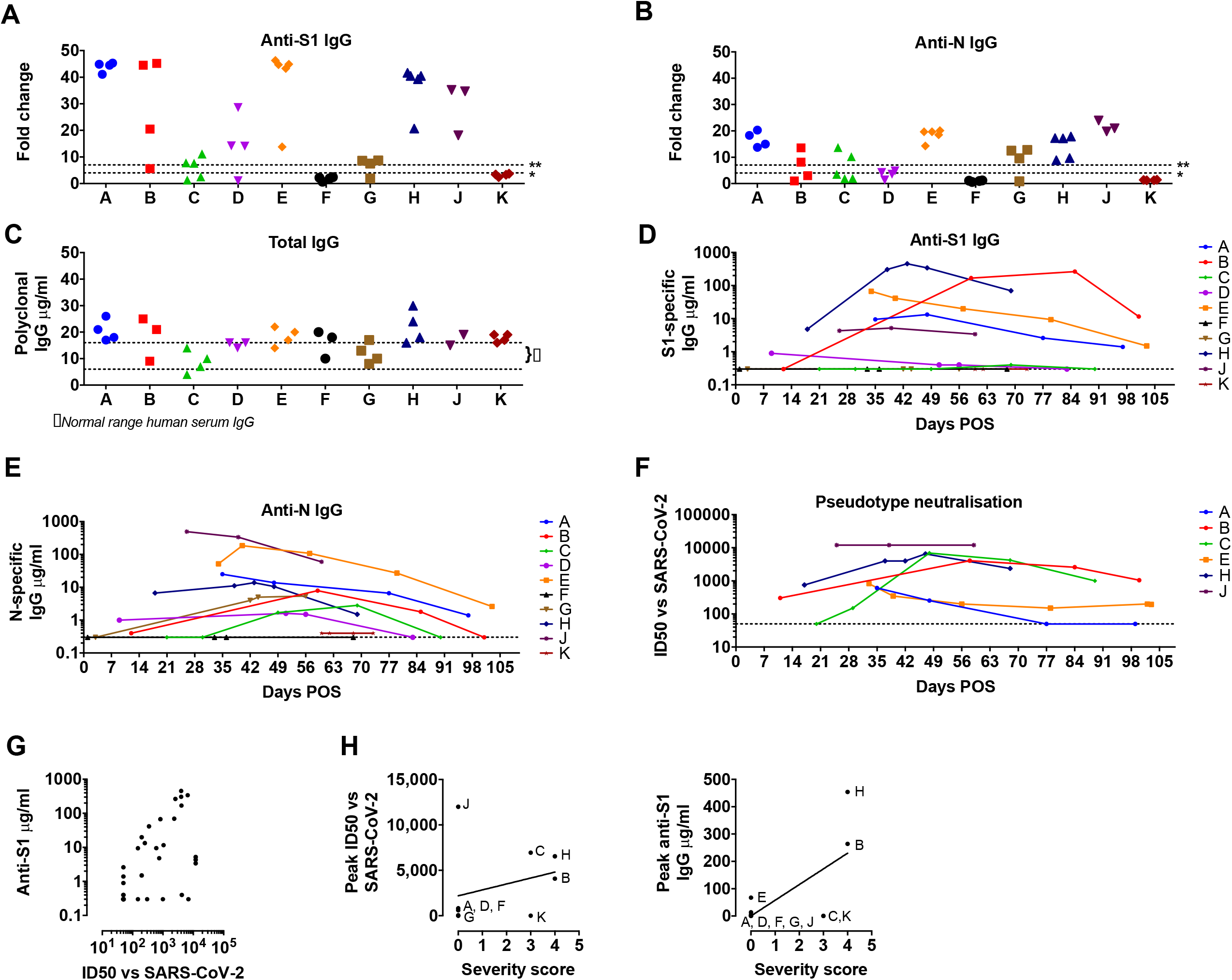
Temporal dynamics and anti-viral function of SARS-CoV-2 antibody responses in haematological malignancy. **A-B**. Serum samples from patients A-K were assayed on ELISA plates pre-coated with S1 (A) or N (B). Antigen production and assay conditions were as previously described^10, 15, 16^ except that all samples were treated with 0.5% NP40 before dilution in ELISA buffer. Absorbance was measured at 405nm and data expressed as fold-change above blank background. The limit of detection (seropositivity) was determined as fold change >4 and is indicated by the dotted line marked *. The dotted line marked ** indicates the limit of quantification in the assay depicted in D-E, determined by the linear range of the standard curve. **C**. ELISA plates were coated overnight at 4°C with goat anti-human F(ab)’2. Serum samples from patients A-K, and commercial polyclonal IgG standard, were titrated in ELISA buffer and added to the ELISA plate. Binding was detected with goat anti-human IgG conjugated to peroxidase and absorbance read at 450 nm. IgG concentrations in serum were calculated based on interpolation from the IgG standard results using a four-parameter logistic (4PL) regression curve fitting model. Dotted lines marked with }* indicate the normal average range for IgG in human serum. **D-E**. Sera supplemented with 0.5% NP40 from patients A-K were diluted in ELISA buffer and then added to a blocked ELISA plates pre-coated with the indicated antigen and three lanes of goat anti-human F(ab)’2 as per (C). Binding was detected with anti-IgG conjugated to alkaline phosphatase and absorbance measured at 405 nm. Antigen-specific IgG concentrations in serum were calculated based on interpolation from the IgG standard results using a four-parameter logistic (4PL) regression curve fitting model. The dotted line indicates the limit of quantification, which is determined by the linear range of the standard curve and higher than the limit of detection in (A-B). **F**. Sequential serum samples from seropositive patients were titrated in duplicate and pre-incubated with luciferase-encoding HIV pseudotyped with the SARS-CoV-2 Spike for 1h prior to the addition of HeLa cells expressing human ACE2 as previously described^17^. ID50 titres were only calculated in GraphPad Prism where at least two data points exhibited >50% neutralisation. ID50 values for each patient are plotted on the y-axis against days POS on the x-axis. **G**. ID50 values plotted against semi-quantitative S1 titres (μg/ml) for each sample. There is a trend to increasing ID50 with higher S1 titres but linear regression does not show a significant correlation (r = 0.06915, p = 0.0925). All data are from at least three independent experiments.

To compare the temporal dynamics of SARS-CoV-2 antibody responses, samples were grouped in seven-day intervals POS (Supplementary Fig. 2). Seroconversion to S appeared to precede N with only two (H and J) seroconverting to both antigens by day 30 (Supplementary Fig. 2 and Supplementary Tables 1, 2). While these conversion rates are similar to those reported in patients without haematological malignancies^15, 16, 17^, the production of anti-N IgG in PHM appears delayed; 50% seroconverted by day 28 compared to 90% of healthy individuals^17^. However, it should be noted that earlier time points from some patients with the strongest responses were not available (Supplementary Fig. 2 and Supplementary Tables 1,2). To further characterise the dynamics of the antibody responses, S1 and N antibodies were measured in a semi-quantitative assay^18^. We observed four patterns of anti-S1 antibody response (Fig. 1D,E); increasing titre consistent with seroconversion (patients B, C, G and H), declining titres suggesting waning responses (patients A, D and E), maintained antibody levels (patient J) and no antibody response (patients F and K). Anti-N responses were broadly similar to those against S1, although differences in antibody titre were apparent. Consistent with other reports, SARS-CoV-2 IgG responses persisted for approximately 100 days after symptom onset (patient B)^17^.

The ability of antibodies to neutralise virus is important for viral clearance and protection from re-infection. To address whether the SARS-CoV-2-specific antibodies generated by PHM are functional and able to inhibit SARS-CoV-2 infection, we used a serum neutralisation assay with pseudotyped SARS-CoV-2, known to correlate with inhibition of infection in live virus assays^19^. We assessed the neutralisation activity of sera from each patient by measuring their capacity to block infection of Hela cells expressing angiotensin converting enzyme 2 (ACE2, the receptor of SARS-CoV-2 Spike) by a luciferase-encoding attenuated HIV that had been pseudo-typed with Spike^17, 19^. Serum was incubated with virus and permissive cells and the 50% inhibitory antibody concentration (ID50) calculated (Fig. 1F). As expected, seronegative patients F and K showed no neutralisation activity. Inhibition of infection was observed in six of the eight seropositive patient samples. Samples from patients D and G showed no neutralisation activity and both had either very low or unquantifiable levels of S1-specific IgG (Fig. 1D and Supplementary Table 1). In general, neutralisation correlated with anti-S1 IgG levels (Fig. 1G); however, some serum samples showed strong neutralisation (ID50 values between ∼1:100 to 10,000) despite low anti-S1 titres (<6 μg/ml), for example all samples from patients C and J neutralised infection despite low anti-S1 IgG levels. This interesting finding could be because; (i) they produced a relatively high proportion of S1-specific IgG with neutralising capacity, (ii) they produced low titres of particularly potent anti-S1 IgG, (iii) they produced neutralising antibodies directed against other viral epitopes, or (iv) they produced non-IgG neutralising antibodies. Of note, patient C (B-ALL), received blinatumomab (anti-CD19 immunotherapy) and also developed severe macrophage activation syndrome requiring Anakinra (anti-IL1), highlighting the complexity in establishing the causes of the variable responses in this cohort and the need to better understand the quality of antibody responses in PHM with different diagnoses and therapies.

Studies of antibody responses to SARS-CoV-2 infection in the wider population show a correlation between clinical severity and the magnitude of antibody responses^16, 17^. We assigned our patients COVID-19 severity scores of 0-4^16^ (Table 1), and those with the strongest anti-S1 IgG and neutralising responses were generally the most severely ill, with scores of 4 (patients B and H). Patients who experienced milder disease (severity score 0, patients A, D and J) produced lower titre S1-specific responses (Fig. 1H). These correlations are not absolute however; patient C had moderately severe disease but low peak IgG titres, and patient J (B-ALL) had mild disease but a strong neutralising response.

To our knowledge, ours is the first longitudinal study of serological responses to SARS-CoV-2 in adult patients undergoing SACT for aggressive haematological malignancies. We found that the majority of patients positive by PCR for SARS-CoV-2 infection develop antibodies to the major SARS-CoV-2 antigens (S1 and N) and their serum blocks viral infection *in vitro*. The magnitude of the antibody response generally correlated with severity of COVID-19, as is reported in the general population^10, 15, 16, 17^; however, there was no obvious correlation between serological response and haematological diagnosis or type or intensity of SACT. Future large, national studies are required to determine the impact of different SACT regimens on the dynamics of immune responses to SARS-CoV-2 and how this may influence patient outcomes.

Although we saw variations in the magnitude and quality of antibody responses in PHM that were similar to those reported for COVID-19 patients without haematological malignancies^15, 17, 19^, we did observe that the time to seroconversion was consistently longer in PHM. This finding may be significant for both case definition and for determining the prevalence of COVID-19 in such patients, but must be confirmed in larger studies, which should include early and frequent PCR testing to determine the precise onset and duration of SARS-CoV-2 infection, and regular serum sampling to capture the emerging antibody response in greater detail. As the development of antibody responses following natural infection or vaccination is likely to be a key parameter in preventing and controlling COVID-19, it is critical to understand these responses in vulnerable groups such as PHM.

## Data Availability

All data will be made available upon reasonable request.

## Author contributions

Designed the study: JO, LEM, CJ

Sample curation: JO, JZ, RG

Performed experiments: LM, LEM, CR-S

Generated reagents: AR, CE, PC

Analysed data: JO, JZ, LM, LEM

Wrote/edited the manuscript: JO, LEM, CJ, LM, JZ, AK, RG

## Acknowledgements

We are extremely grateful to all the patients who participated in this study and the NHS staff that provided their clinical care. We would like to thank, Leo James and Jakub Luptak (LMB) for the provision of the plasmid encoding the N protein, and James E Voss (TSRI) for providing the Hela-ACE2. LEM and LM are supported by an MRC Career Development Award (MR/R008698/1) to LEM. CJ is supported by a Wellcome Trust Investigator award (108079/Z/15/Z). R.G. acknowledges funding from Cure Cancer@ UCL.

## Ethical statement

All clinical information was recorded and blood samples taken as routine standard of care. Patients were consented to allow any excess serum to be stored and used as part of the “UCL Biobank for Studying Health and Disease – Haematology Project”, reference no NC10.13, approved by the Leeds (East) Research Ethics Committee, UK.

## SUPPLEMENTARY INFORMATION

**Supplementary figure 1: Duration of SARS-CoV-2 infection by PCR testing**.

**Supplementary figure 2: IgG and IgM responses in this cohort**

**Supplementary table 1: Anti-S1 IgG responses**

**Supplementary table 2: Anti-N IgG responses**

**Supplementary table 3: Anti-S1 IgM responses**

**Supplementary figure 1:**
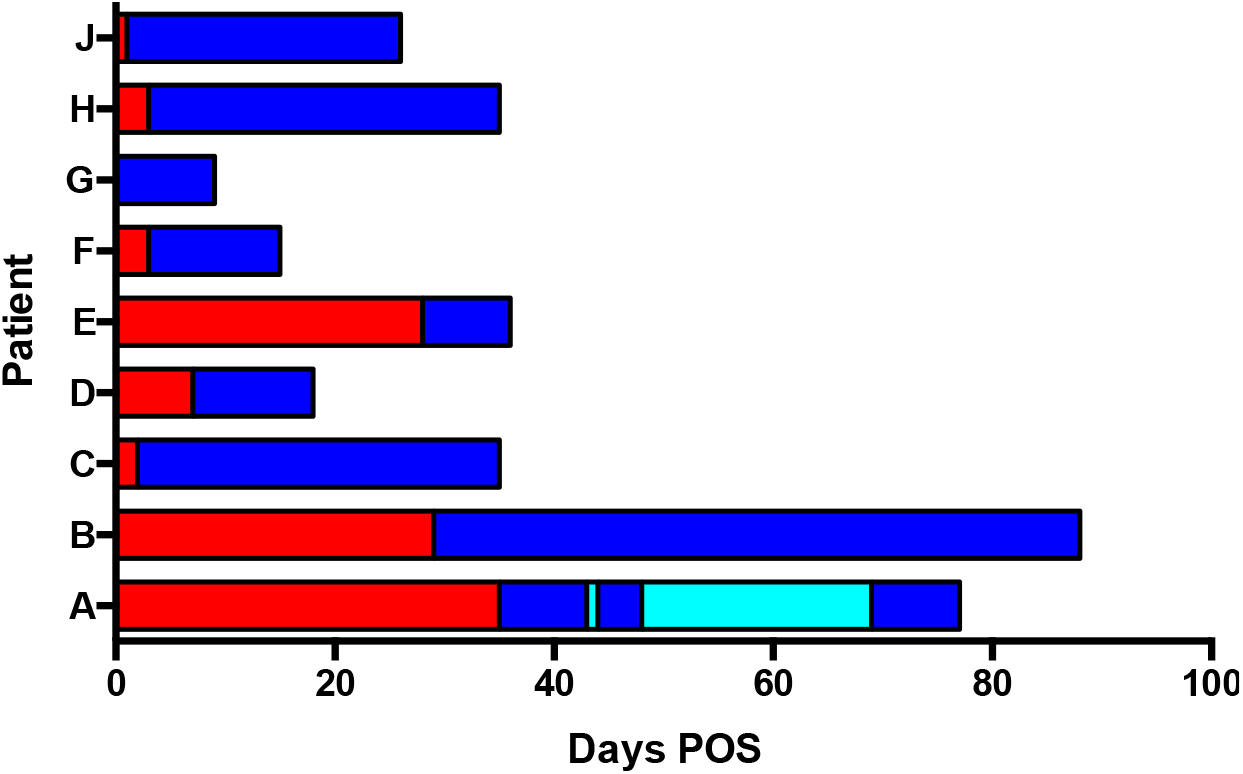
Duration of SARS-CoV-2 infection by PCR testing. Graph showing the time from COVID-19 POS to confirmed SARS-COV-2 infection by PCR of nasopharyngeal samples (red) and the number of days each patient tested positive for SARS-CoV-2 by PCR (blue). The duration of any PCR negative periods is shown in cyan. All patients tested negative by PCR surveillance. Patient K was not SARS-CoV-2 positive by PCR and is not shown.

**Supplementary figure 2:**
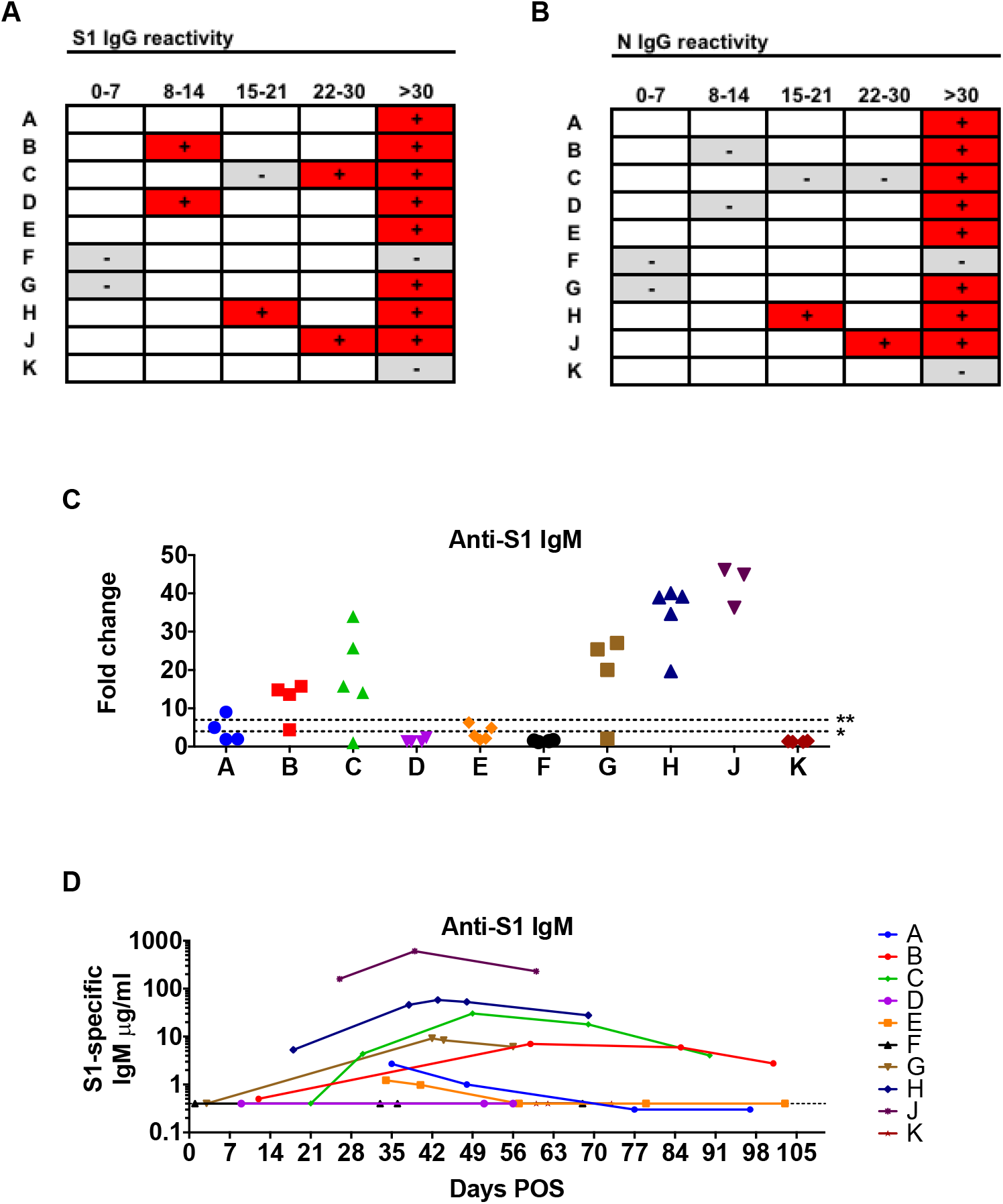
IgG and IgM responses in this cohort. **A-B**. S1 (A) and N (B) antigen IgG seroconversion. Serum samples that gave a signal >4-fold above background in the seropositivity ELISA are marked with a “+” sign and colour-coded red. Those that gave a signal <4-fold above background are marked with a “-” sign and colour-coded grey. Blank boxes indicate that no serum sample was taken from the patient (indicated in the left-hand column) at that time interval (indicated in the row titles across the top). **C**. Serum samples from patients A-K were diluted to 1:50 in ELISA buffer (PBS, 5% milk, 0.05% Tween 20) prior to addition to an ELISA plate pre-coated with S1 (A). Antigen production and assay conditions were as previously described^8,13,14^ except that all samples were treated with 0.5% NP40 before dilution in ELISA buffer and binding was detected using anti-IgM conjugated to alkaline phosphatase (IgM-AP). Absorbance was measured at 405nm and data expressed as fold-change above blank background. The limit of detection (seropositivity) was determined as fold change >4 and is indicated by the dotted line marked. **D**. Serum supplemented with 0.5% NP40 from patients A-K was diluted in ELISA buffer and then added to a blocked ELISA plate pre-coated with the indicated antigen and three lanes of goat anti-human F(ab)’2 as per (C) to facilitate the generation of the standard curve. Diluted serum samples were incubated on the assay plate for 2 h. Plates were washed, incubated with goat anti-human IgM-AP diluted in ELISA buffer, washed and developed using alkaline phosphatase with absorbance measured at 405 nm. Antigen-specific IgM concentrations in serum were then calculated based on interpolation from the IgM standard results using a four-parameter logistic (4PL) regression curve fitting model. The dotted line indicates the limit of quantification in which is determined by the linear range of the standard curve and higher than the limit of detection in C.

**Supplementary table 1:**
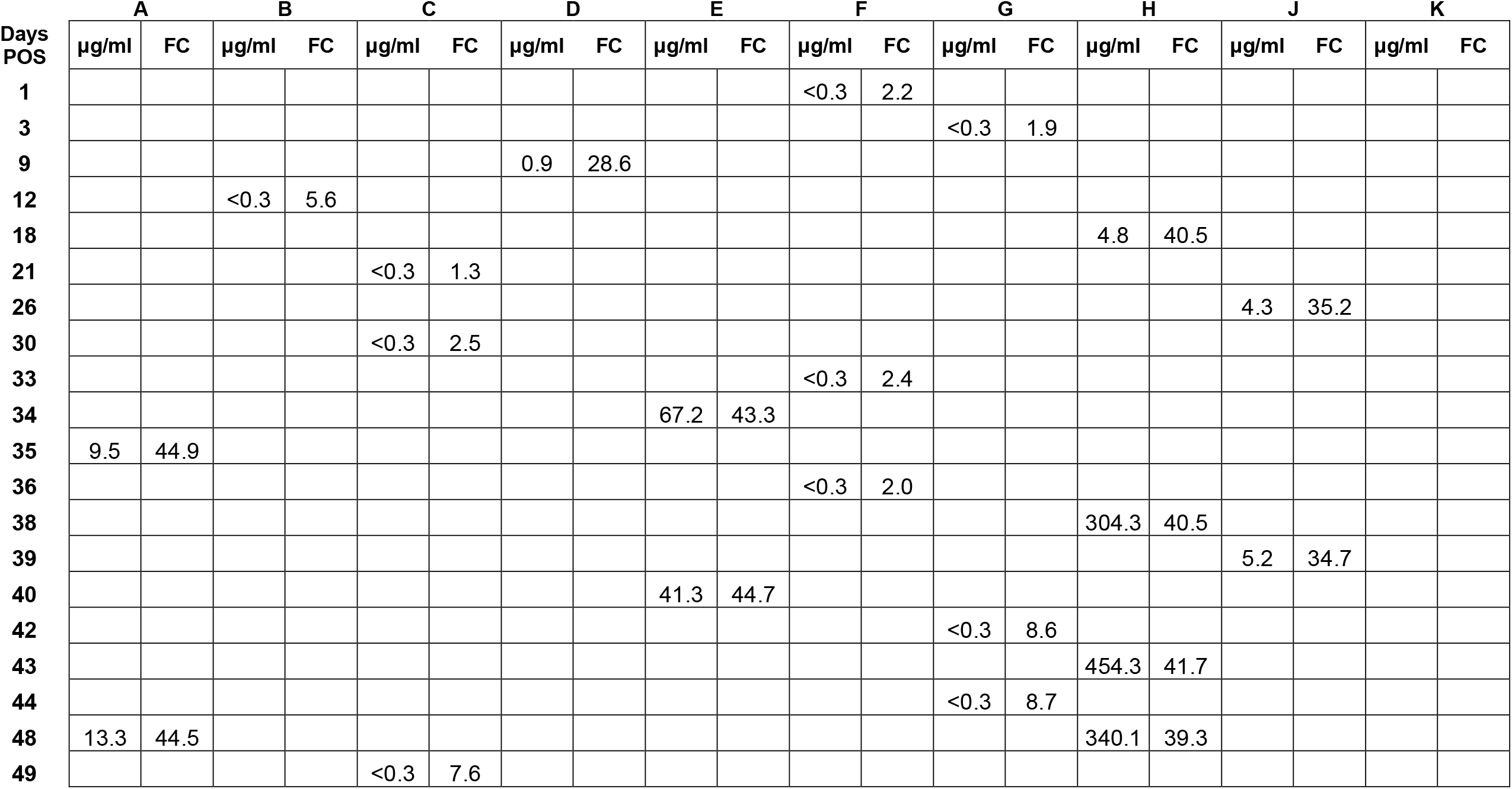
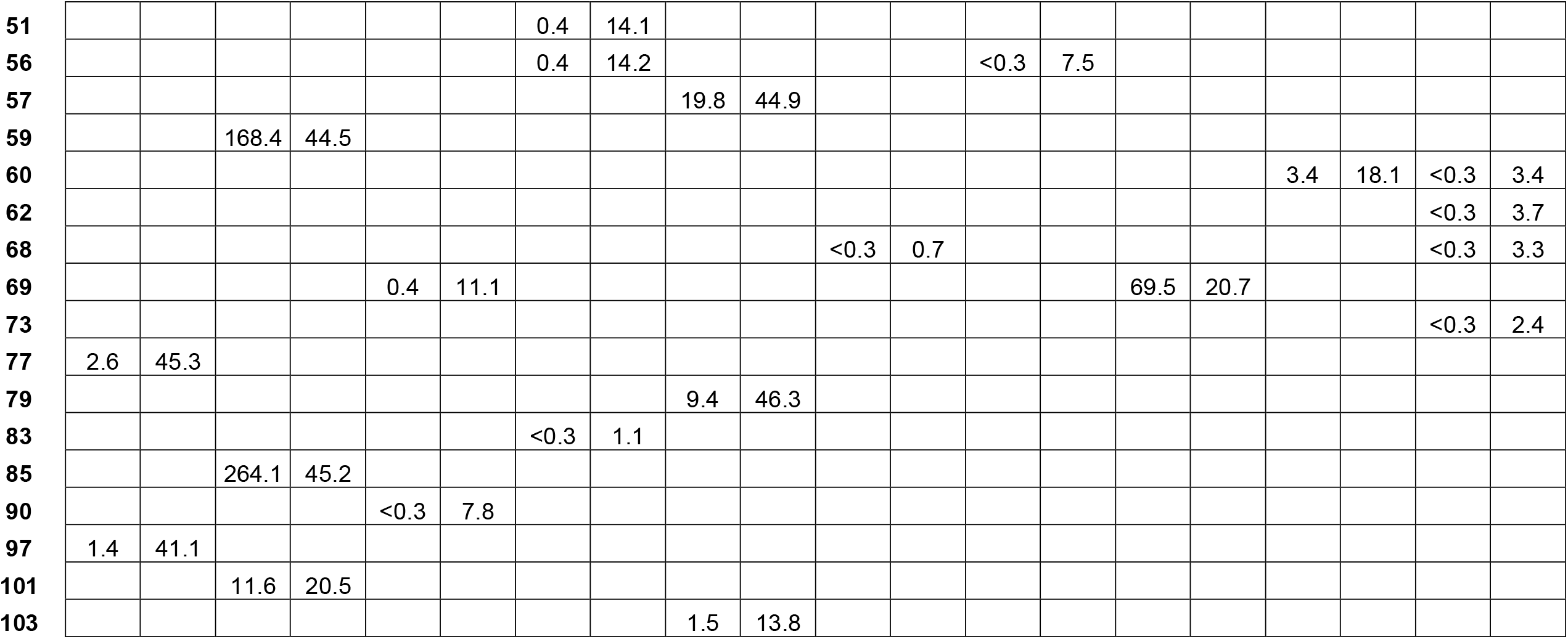
Anti-S1 IgG responses. Semi-quantitative titres in μg/ml and qualitative binding in fold-change (FC) above average blank absorbance are shown for each patient, tabulated by the day post symptom onset (POS) on which the sample was taken.

**Supplementary table 2:**
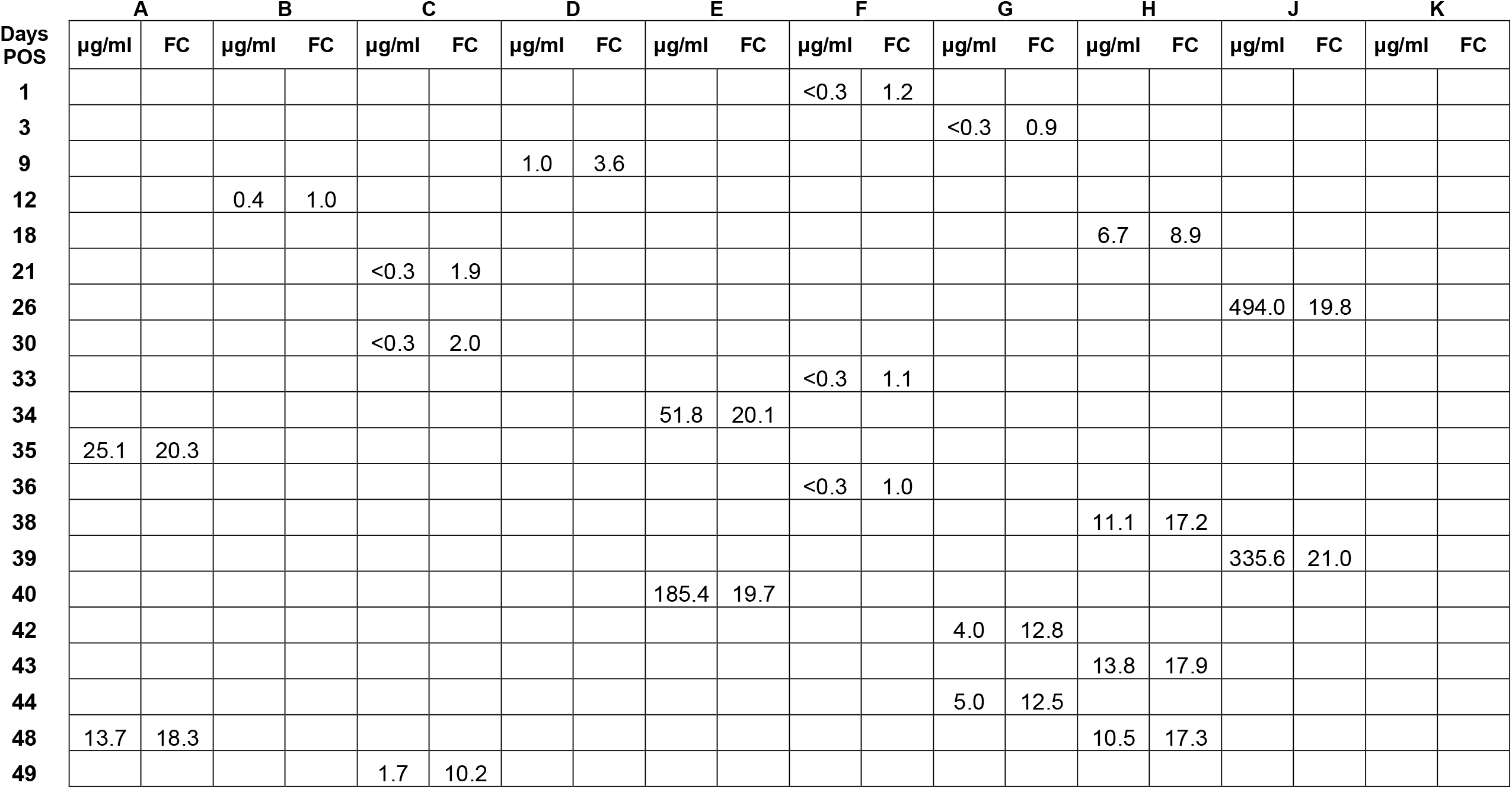
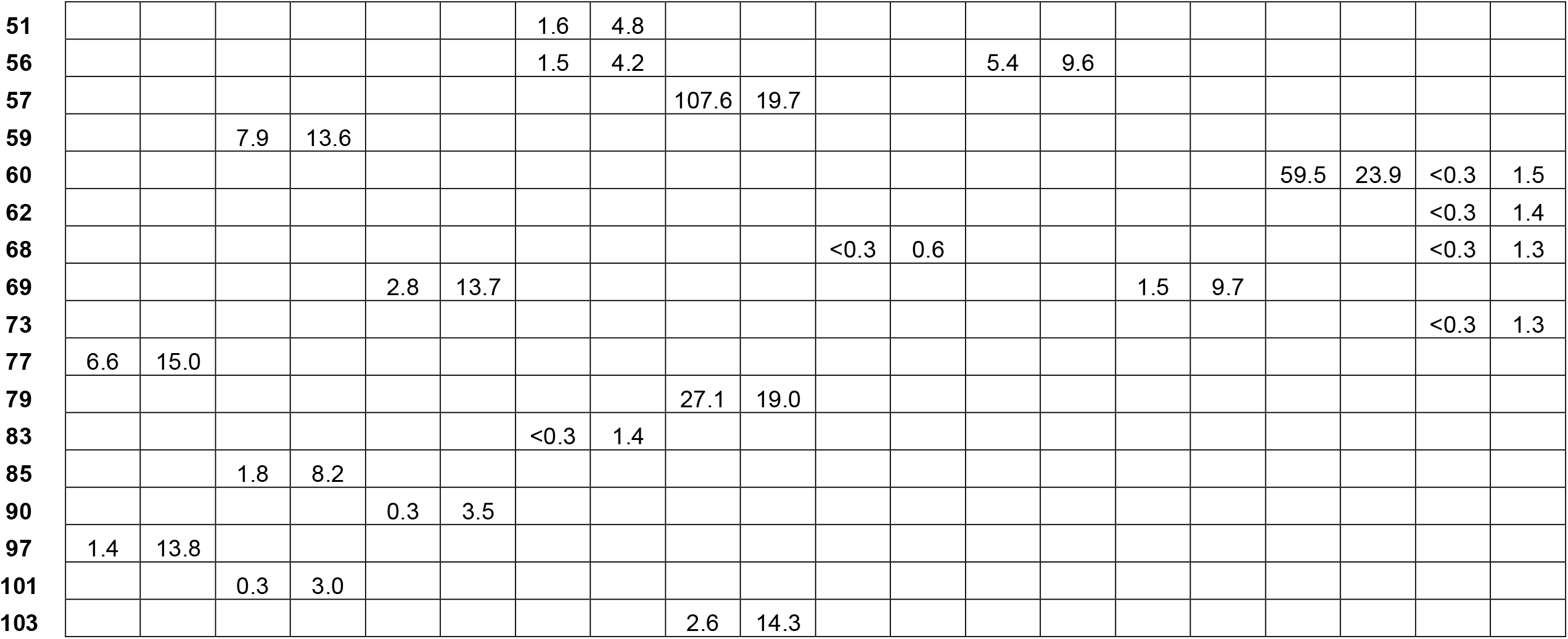
Anti-N IgG responses. Semi-quantitative titres in μg/ml and qualitative binding in fold-change (FC) above average blank absorbance are shown for each patient, tabulated by the day post symptom onset (POS) on which the sample was taken.

**Supplementary table 3:**
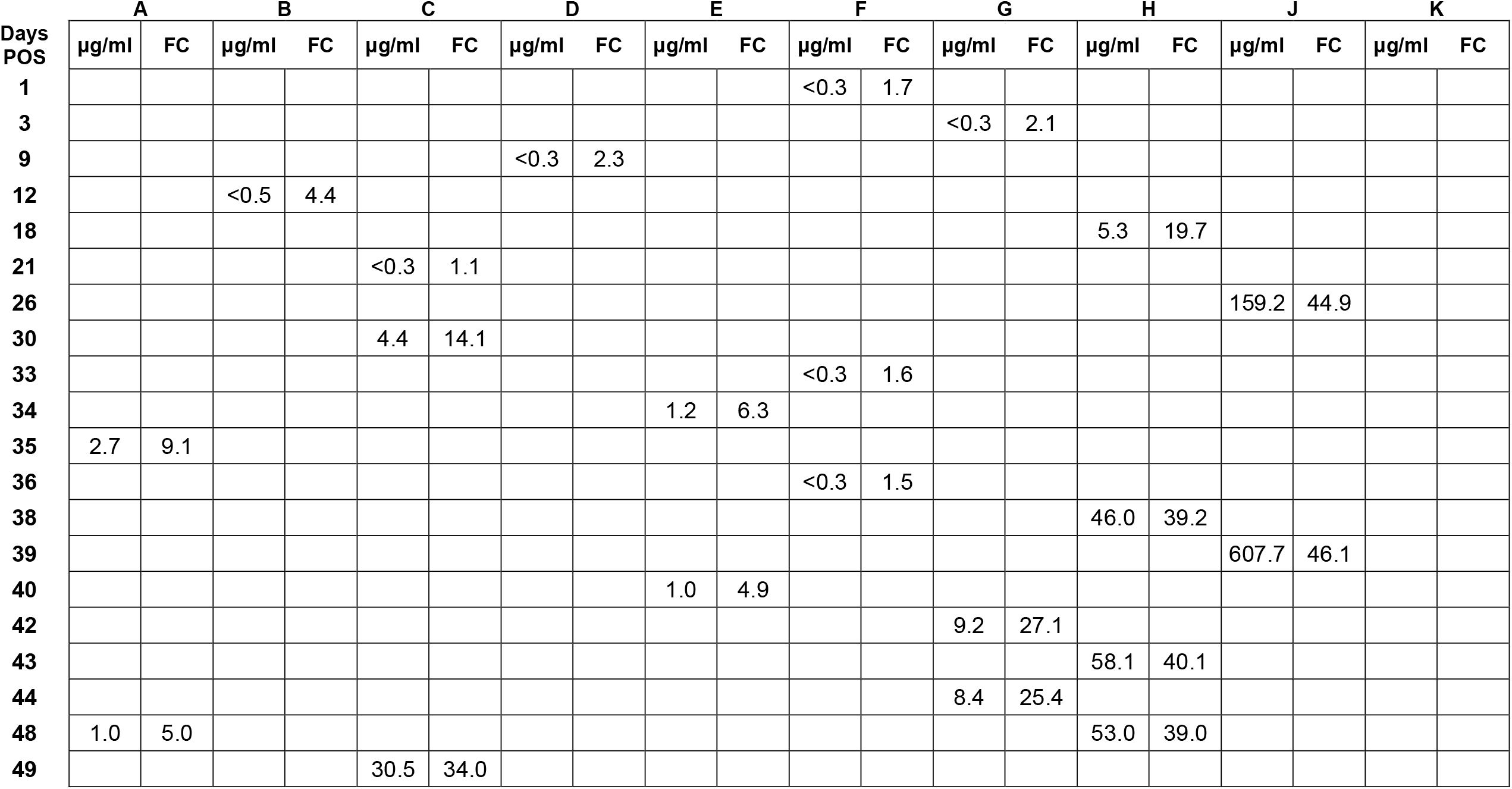
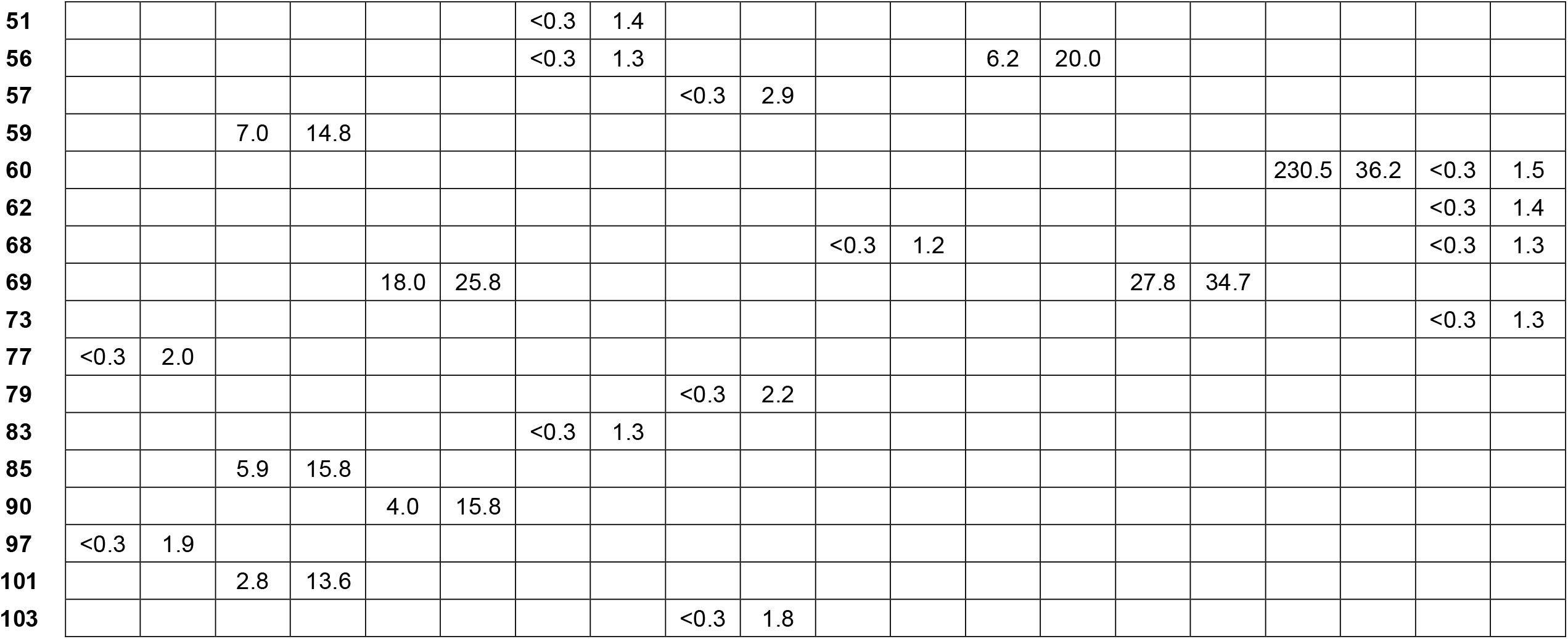
Anti-S1 IgM responses. Semi-quantitative titres in μg/ml and qualitative binding in fold-change (FC) above average blank absorbance are shown for each patient, tabulated by the day post symptom onset (POS) on which the sample was taken.

